# Gender-specific associations of adverse childhood experiences (ACEs) and schizotypal traits – an observational study in healthy young adults

**DOI:** 10.1101/2024.07.08.24310072

**Authors:** Kirchhoff Christina, Riedl David, Rothmund Maria-Sophie, Hüfner Katharina, Scantamburlo Gabrielle, Scholtes Felix, Brandenberg Marius, Steiner Anna, Dannecker Noemi, Surbeck Werner, Homan Philipp

**Affiliations:** University Hospital of Psychiatry II, Department of Psychiatry, Psychotherapy Psychosomatics and Medical Psychology, Medical University of Innsbruck, Innsbruck, Austria; Ludwig Boltzmann Institute for Rehabilitation Research, Vienna, Austria; Department of Psychiatry, CHU of Liege; Psychoneuroendocrinology Unit, University of Liege, Belgium; Service de Neuroanatomie, Department of Preclinical Science, University of Liege, Belgium; Service de Neurochirurgie, Department of Surgery, Centre Hospitalier Universitaire de Liège, Belgium; Department of Adult Psychiatry and Psychotherapy, Psychiatric Hospital, University of Zurich, Zurich, Switzerland; Neuroscience Center Zurich, University of Zurich and ETH Zurich, Zurich, Switzerland

**Keywords:** gender, adverse childhood experiences, maltreatment, schizotypy, healthy young adults

## Abstract

**Background and Hypotheses:** Schizotypy is a complex model containing a broad spectrum of personality traits that can be observed in the general population as well as in psychiatric patients. There is compelling evidence that Adverse Childhood Experiences (ACEs) are correlated with schizotypal traits in healthy individuals. We hypothesize that associations between specific forms of abuse and distinct schizotypal traits will demonstrate gender-specific differences.

**Study Design:** The present study relies on a dataset designed and collected for the VELAS-study (VELAS: Ventral language stream in schizophrenia with regard to semantic and visuo-spatial processing anomalies) in Zurich, Switzerland. Young adults completed an online questionnaire which included the Childhood Trauma Questionnaire (CTQ) and the Oxford-Liverpool Inventory of Feelings and Experiences (O-LIFE). To test individual associations, gender-specific linear regression models were calculated.

**Study Results:** A total of 597 healthy young adults completed the online questionnaire. Gender-specific linear regression models revealed strong associations of emotional abuse with all schizotypal traits in both sexes with stronger effect sizes for male subjects. In men, sexual abuse was associated with Unusual Experiences, while in women it was associated with Disorganisation. Emotional neglect showed an association with Introvertive Anhedonia in both genders, with stronger effect sizes for male than female participants. Physical neglect exhibited an association with Introvertive Anhedonia solely in male subjects.

**Conclusions:** Our gender-specific results give a deeper insight into associations of ACEs with schizotypal traits and serve as a puzzle piece in understanding risk constellations in the development of schizophrenia spectrum disorders.

## Introduction

There is compelling evidence that childhood trauma contributes to the occurrence of psychosis and schizophrenia spectrum disorders (Afifi et al., 2011; Beards et al., 2013; Lobbestael, Arntz, & Bernstein, 2010; Morgan et al., 2014). Schizotypy is widely acknowledged as a cluster of personality traits. These encompass a range of emotional, cognitive, and perceptual experiences commonly observed in schizophrenia spectrum disorders. Schizotypy thus serves as a foundational framework for the study of susceptibility to these disorders (Claridge, 1997; Kwapil & Barrantes-Vidal, 2014; Meehl, 1962). This construct comprises a complex model that includes ‘positive,’ ‘negative,’ and ‘disorganized’ dimensions, which do not inherently imply a pathogenic state but may encompass favorable and protective aspects (Flückiger et al., 2019; Christine Mohr & Gordon Claridge, 2015).

Claridge et al introduced a fully dimensional model of schizotypy, highlighting schizotypy as a broad spectrum of personality characteristics, both healthy and unhealthy, that can be observed in the general population as well as in psychiatric patients (Claridge, 1997). In line with this dimensional approach, the Oxford-Liverpool Inventory of Feelings and Experiences (O-LIFE) was developed, based on four scales: Unusual Experiences, including aberrant perceptions and magical thinking, Cognitive Disorganisation, including attentional difficulties, social anxiety, odd behavior and odd speech, Introvertive Anhedonia, including a lack of enjoyment from social and physical sources of pleasure and lack of intimacy, and Impulsive Nonconformity, including impulsive, eccentric and anti-social behavior and lack of self-control (Mason, Claridge, & Jackson, 1995). While the first three scales address the dimensions of the three factor model of schizophrenic symptoms (Vollema & Hoijtink, 2000), the fourth scale (Impulsive Nonconformity) was included later to account for additional personality dimensions and symptoms related to bipolar disorders. Consistent data indicates that younger individuals tend to report higher scores on all scales except for Introvertive Anhedonia (Mason & Claridge, 2006).

Numerous original studies (e.g., Gong, Wang, Lui, Cheung, & Chan, 2017; Sheinbaum, Kwapil, & Barrantes-Vidal, 2014; Mike Startup, 1999) and two recent meta-analyses (Toutountzidis, Gale, Irvine, Sharma, & Laws, 2022; Velikonja, Fisher, Mason, & Johnson, 2015) have demonstrated a correlation between Adverse Childhood Experiences (ACEs) and schizotypal traits in healthy individuals and suggest a dose-response relationship. All forms of ACEs, namely emotional, physical, and sexual abuse, as well as emotional and physical neglect, are linked to schizotypal traits, where emotional abuse demonstrates the most substantial effects (Alemany et al., 2011; Ered & Ellman, 2019; Toutountzidis et al., 2022). Recently, the ENIGMA working group suggested that childhood trauma severity moderates relationships between schizotypy and brain morphology in psychiatrically healthy adults (Quidé et al., 2024). In addition, meta-regression analyses have revealed that the connection between physical abuse and schizotypy is more pronounced in samples with a higher proportion of women and younger participants (Toutountzidis et al., 2022). Berenbaum et al demonstrated that the association of childhood maltreatment and schizotypal traits is stronger in men than women. In women, schizotypal symptoms were more strongly associated with Criterion A of posttraumatic stress disorder than in men (Berenbaum, Valera, & Kerns, 2003). It is worth noting that gender differences in symptoms and the course of patients with schizophrenia are well-documented (Giordano, Bucci, Mucci, Pezzella, & Galderisi, 2021; Li, Zhou, & Yi, 2022).

While numerous studies have explored the connection between ACEs and schizotypal traits, only a limited number of studies in healthy individuals have focused on gender differences and a nuanced examination of how different ACEs might be associated with specific schizotypal traits. Further limitations of published data are due to the use of schizotypy scores that have been designed for clinical purposes, especially the detection of schizotypal personality disorder, but not to examine mild schizotypal traits in healthy individuals. To address this gap, and building on the review by Toutountzidis et al, we hypothesized a) a dose-dependent association of ACEs with schizotypal traits with the strongest effect for emotional abuse b) gender-specific differences of associations between ACEs and distinct schizotypal traits.

## Methods

### Sample and data

The present study is based on a dataset designed and collected for the recruitment of possible subjects for the VELAS-study (VELAS: Ventral language stream in schizophrenia with regard to semantic and visuo-spatial processing anomalies) at the Individual Differences in Psychosis (IDP) Lab at the University Hospital of Psychiatry in Zurich, Switzerland. Due to different research questions, the following criteria had to be met for inclusion in the VELAS study: subjects had to be between 18 and 35 years old, free from preexisting psychiatric or neurological diseases and right-handed. The dataset is based on an online assessment distributed over various platforms and noticeboards of Swiss universities and colleges, Swiss public platforms, a mailing list of students of psychology, tweets by the research team and direct contact of different schools and educational institutions of various levels. The dataset includes demographic items, the German version of the O-LIFE and the German version of the Childhood Trauma Questionnaire (CTQ).

### Measures

The CTQ is a retrospective self-report questionnaire to assess the nature and the intensity of early traumatic experiences (Bernstein et al., 1994). It consists of 25 items which assess various forms of trauma, including physical and sexual abuse, emotional neglect, emotional abuse, and physical neglect, as well as a three-item minimisation/denial scale indicating the potential underreporting of maltreatment. The items can be scored from 1 (never true) to 5 (very often true), leading to a total score in each subscale ranging from 5 to 25 points. Severity levels of abuse or neglect can be classified for each subscale using defined cutoff scores, categorizing them as none or minimal, low to moderate, moderate to severe, or severe to extreme. The German version of the CTQ has demonstrated strong internal consistencies, factorial validity, and both convergent and discriminant validity (Klinitzke, Romppel, Häuser, Brähler, & Glaesmer, 2012). The O-LIFE is a widely adopted, multidimensional tool for assessing schizotypy. (Mason et al., 1995), comprising 104 Yes/No items. Good psychometric properties for the German version of the O-LIFE were reported (Phillip Grant et al., 2013).

### Statistical procedures

Demographics for the sample are presented with means and standard deviations (SD). Patients with missing items in CTQ or O-LIFE were excluded from the analysis. To display distribution of ACEs and schizotypal traits, data was grouped based on reference data. Severity of ACEs was grouped into none, slight, moderate and severe based on the cut-offs proposed by (Hauser, Schmutzer, Brahler, & Glaesmer, 2011). O-life subscales were compared to normative data published by (Mason & Claridge, 2006) and were grouped into low (<25^th^ percentile), moderate (25-75^th^ percentile) and high (>25^th^ percentile) scores.

The association of CTQ subscales and schizotypal traits with age and gender was tested with independent sample t-tests and Pearson correlation coefficients. The association of ACEs on the overall schizotypal trait was tested with a stepwise linear regression model: in step (1) sociodemographic variables (age, gender, family history of schizophrenia) were added to the model and in step (2) the CTQ subscales. Due to the relatively low number of cases, physical abuse was not used in the following models. The model tested the significant increase of explained variance in each step. Durbin-Watson and VIF statistics were calculated to test for autocorrelation and multicolinearity, with Durbin-Watson values between 1.5 to 2.5 and VIF <5 considered acceptable.

To test the relationships of different ACE types with the four specific schizotypal traits (Unusual Experiences, Cognitive Disorganization, Introvertive Anhedonia, Impulsive Nonconformity) gender specific linear regression models were calculated with the ACE types as independent variables and the respective O-LIFE scale as dependent variable. P-values <0.05 were considered statistically significant. All calculations were performed with IBM SPSS (v22.0).

## Results

A total of 613 participants completed the assessment for the VELAS study. Of these, n = 16 participants (2.6 %) were excluded because of missing data in the CTQ. The remaining n = 597 participants were included in the present analysis. Participants’ mean age was 24.5 years and the majority was female (78.6%). For more details on sociodemographic characteristics see table 1.

**Table 1:** Sociodemographic and clinical characterstics

|  | N | % |
| --- | --- | --- |
| Sex |  |  |
| female | 469 | 78.6% |
| male | 128 | 21.4% |
| Age (M=24.5; SD=4.2) |  |  |
| <25 years | 341 | 57.1% |
| 25-30 years | 192 | 32.2% |
| >30 years | 64 | 10.7% |
| Highest level of education |  |  |
| no high-school diploma | 143 | 24.0% |
| high-school diploma and higher education | 453 | 75.9% |
| missing | 1 | 0.2% |
| Family history of schizophrenia |  |  |
| None | 560 | 93.8% |
| 1<sup>st</sup> degree | 14 | 2.3% |
| 2<sup>nd</sup> degree | 23 | 3.9% |

### Prevalence of ACEs

A total of 227 (38.0 %) participants reported no ACEs, while 334 (55.9 %) had experienced 1-3 ACEs and the remaining 36 (6.0 %) four or more ACEs (i.e., polytraumatized persons). Most frequent form of ACE was emotional abuse (n = 209, 35.0%), followed by emotional neglect (n = 187, 31.3%), physical neglect (n = 146, 24.5%) and sexual abuse (n = 136, 22.8%), while physical abuse was reported least frequently (n = 49, 8.2%).

With regard to severity of ACEs, most frequently reported moderate to severe abuse was found for emotional abuse (n = 64, 10.7%) and sexual abuse (n = 64, 10.7%). Gender specific analyses of abuse severity showed no significant differences neither for emotional (χ^2^=5.68, p=.13) and physical abuse (χ^2^=3.12, p=.37), nor for emotional (χ^2^=3.04, p=.39) or physical neglect (χ^2^=3.76, p=.29). However, sexual abuse was found more frequently among women than men (χ^2^=16.70, p< .001), with women reporting moderate to severe forms of sexual abuse almost three times as often as men (12.4% vs. 4.7%). The overall number of reported ACEs however was neither associated to age (r = .07, p = .10) nor gender (t = 1.40, p = .16). The prevalence of ACEs and reported severity per gender are graphically displayed in figure 1.

**Figure 1.**
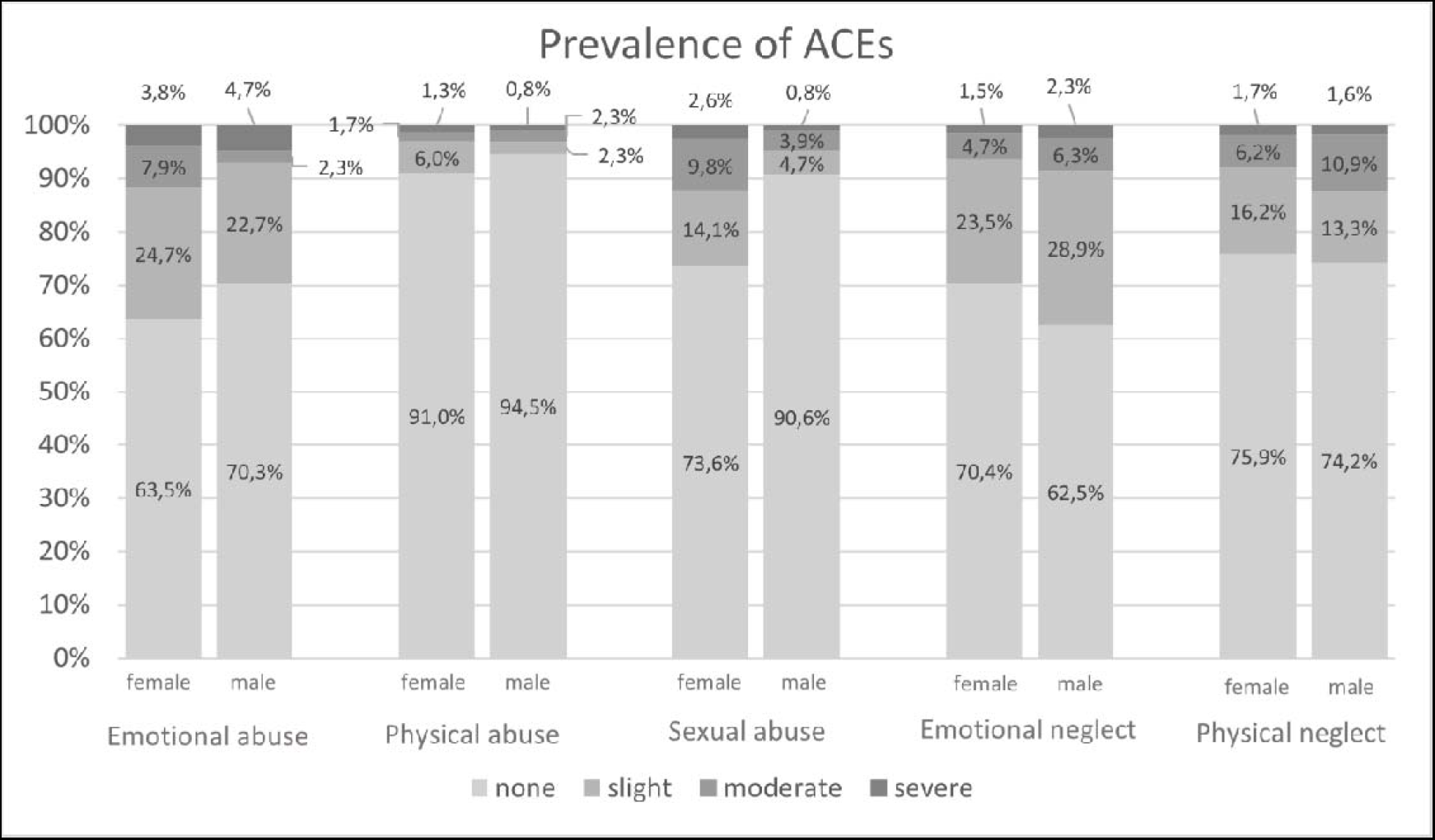
Prevalence of adverse childhood experiences (ACEs), stratified by gender

### Prevalence of schizotypal traits

A majority of participants reported either low or moderate scores on all four assessed schizotypal traits. However, 19.8% (n = 118) of the sample showed higher scores (i.e., above the 75^th^ percentile of the reference population). The greatest percentage of high scores were found for the subscale Introvertive Anhedonia (i.e., ‘negative symptoms’), while for the other three subscales, the number of high scores was comparably low. Gender specific analyses of the distribution among all four assessed schizotypal traits showed no statistically significant difference for Introvertive Anhedonia (χ^2^=2.77, p=.29). However, male participants reported significantly lower scores regarding Cognitive Disorganization (χ^2^=25.39, p<.001) and Unusual Experiences (χ^2^=6.43, p=.040), while women reported low scores for Impulsive Nonconformity (χ^2^=12.86, p=.002). An overview is provided in figure 2.

**Figure 2.**
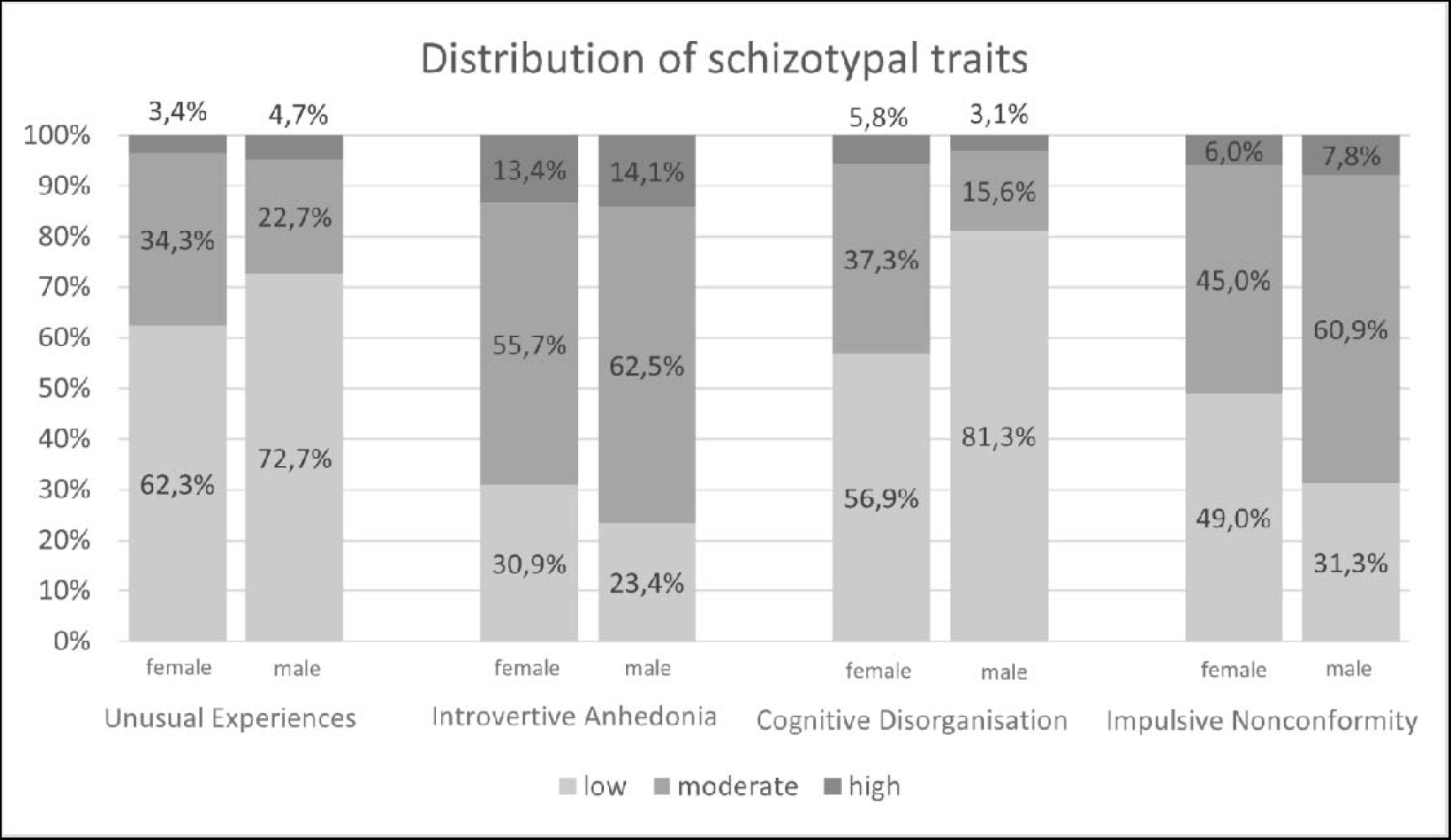
Distribution of schizotypal traits, stratified by gender

### Association of schizotypal traits and ACEs

Higher levels of ACEs were associated with more pronounced overall schizotypal traits (r = .38, p < .001). To evaluate the unique association of emotional and sexual abuse, as well as emotional and physical neglect, with overall schizotypal traits in addition to the assessed sociodemographic variables, a stepwise linear regression model was calculated. The model was statistically significant (F(6, 596)=32.948, p < .001) and no auto-correlation (Durbin-Watson=1.92) or multicollinearity (VIF=1.01-2.01) was observed. Two of the three sociodemographic variables in step 1 (age and family history of schizophrenia) were statistically significant and explained 2.8% of the variance in the O-LIFE total score. When the CTQ subscales (except physical abuse) were added in step 2, a substantial increase of the explained variance to 22.6% (p<.001) was observed. Emotional and sexual abuse were significantly associated with higher overall O-LIFE scores, while no such association was observed for emotional or physical neglect. For details see table 2.

**Table 2:** Hierarchical multiple linear regression model for the association of sociodemographic variables and adverse childhood experiences (CTQ) with schizotypical traits (O-LIFE total score)

|  | Step 1: |  | Step 2: |  |
| --- | --- | --- | --- | --- |
|  | sociodemographic factors |  | ACEs |  |
| | $\beta$ | p value | $\beta$ | p value |
| Age | -.11 | .007 | -.11 | .003 |
| Gender (0=female) | .01 | .85 | .03 | .47 |
| Family history of schizophrenia (0=no, 1=yes) | .14 | .001 | .09 | .014 |
| Emotional abuse | -- | -- | .32 | <.001 |
| Sexual abuse | -- | -- | .09 | .024 |
| Emotional neglect | -- | -- | .08 | .13 |
| Physical neglect | -- | -- | .08 | .07 |
| R<sup>2</sup> (Sig. model) | 2.8% | .001 | 22.6% | <.001 |
| $\Delta R^2$ (Sig. of $\Delta R^2$) | -- | -- | 19.8% | <.001 |
| $\beta$ = standardized coefficient; ACE= Adverse Childhood Experience; | | | | |

### Dose-dependent relationship of ACEs and schizotypal traits

To test the dose-dependent hypothesis, the mean O-LIFE total score for patients with 0 ACEs, 1-3 ACEs and 4+ ACEs were compared. In the model, increasing numbers of ACEs were clearly associated with a higher O-LIFE total score with a large effect size (F(2, 596)=43.327, p < .001, η^2^ = .127), as shown in Figure 3. Patients without ACEs reported a mean O-LIFE total score of 15.2 (SD=7.9) points, while for patients with 1-3 ACEs the score was 21.8 (SD=11.8) points and with 4+ ACEs 30.3 (SD=15.9) points.

**Figure 3.**
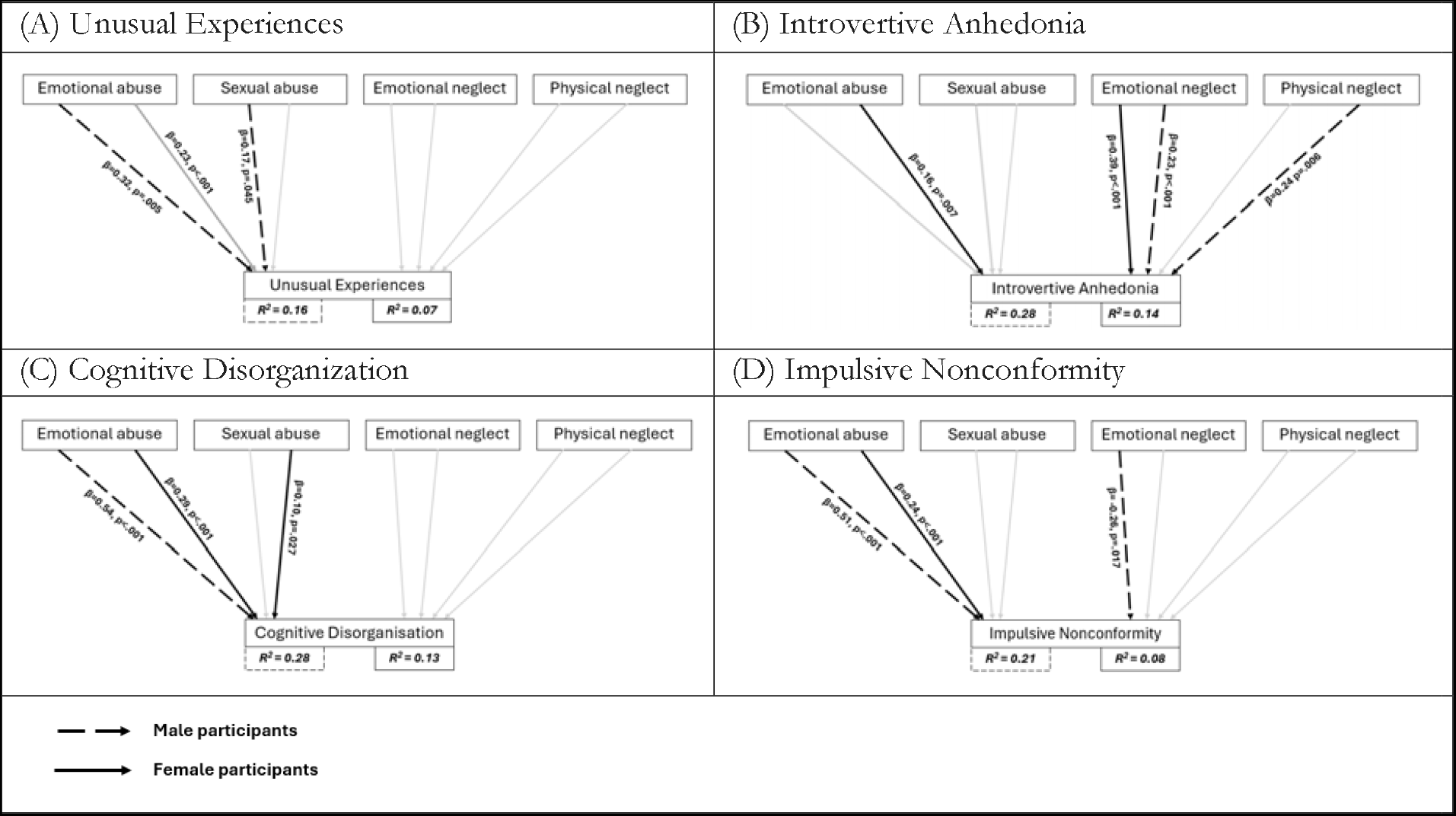
Linear regression models for the association of assessed adverse childhood experiences (CTQ subscales) and the four schizotypal traits (O-LIFE subscales) in the female (n=469) and male sample. Solid lines represent beta-weights and p-values for female participants and dashed lines for male participants. Grey lines represent non-significant paths.

### Specific associations of ACEs with schizotypal traits

To investigate the specific association of individual ACEs on the four schizotypal traits, gender-specific linear regression models were calculated. For details, see figure 3. The model for Unusual Experiences was statistically significant for both genders (female: F(4, 468)=8.789, p < .001; male: F(4,127)=5.874, p < .001) and no auto-correlation (female: Durbin-Watson=1.92; male: Durbin-Watson=2.08) or multicollinearity (female: VIF=1.09–2.02; male: VIF=1.02–1.88) was observed. Emotional abuse significantly predicted Unusual Experiences in both female (β=0.23, p<.001) and male participants (β=0.32, p=.005) while an additional association of sexual abuse (β=0.17, p=.045) was only observed in male participants. The ACE subscales explained a substantially higher proportion of the variance in male than in female participants (16% vs. 7%).

The model for Introvertive Anhedonia was also statistically significant for both genders (female: F(4, 468)=18.895, p < .001; male: F(4,127)=12.002, p < .001) and no auto-correlation (female: Durbin-Watson=1.91; male: Durbin-Watson=1.83) or multicollinearity (female: VIF=1.09–2.02; male: VIF=1.02–1.88) was observed. Emotional abuse (β=0.16, p=.007) and emotional neglect (β=0.39, p<.001) significantly predicted introvertive anhedonia in women and explained 14% of the variance, while emotional neglect (β=0.23, p<.001) and physical neglect (β=0.24 p=.006) were significant predictors in the male sample, explaining 28% of the variance.

The model for Cognitive Disorganization was also statistically significant for both genders (female: F(4, 468)=17.362, p < .001; male: F(4,127)=11.826, p < .001) and no auto-correlation (female: Durbin-Watson=1.91; male: Durbin-Watson=2.07) or multicollinearity (female: VIF=1.02–2.02; male: VIF=1.02–1.88) was observed. Emotional abuse significantly predicted Cognitive Disorganization in both female (β=0.29, p<.001) and male participants (β=0.54, p<.001) while an additional association of sexual abuse (β=0.10, p=.027) was only observed in female participants. The ACE subscales explained a substantially higher proportion of the variance in male than in female participants (28% vs. 13%).

Finally, the model for Impulsive Nonconformity was was also statistically significant for both genders (female: F(4, 468)=10.382, p < .001; male: F(4,127)=8.534, p < .001) and no auto-correlation (female: Durbin-Watson=2.01; male: Durbin-Watson=1.96) or multicollinearity (female: VIF=1.09–2.02; male: VIF=1.02–1.88) was observed. Emotional abuse significantly predicted impulsive non-conformity in both female (β=0.24, p<.001) and male participants (β=0.51, p<.001) while an additional association of emotional neglect (β= -0.26, p=.017) was only observed in male participants. As with the other O-LIFE scales, a substantially higher proportion of variance was explained by the ACE subscales for male than for female participants (21% vs. 8%).

## Discussion

### Overall findings and the association of ACEs with schizotypy total scores

The aim of the present study was to investigate gender-specific associations of different ACEs with schizotypal traits in healthy young adults. Our results mainly confirm the characteristic gender-specific distribution of schizotypal traits in healthy individuals and underline the dose-dependent association of ACEs with schizotypal traits.

In line with the findings of Mason et al, female participants in our sample expressed significantly more Cognitive Disorganisation but less Impulsive Nonconformity than men (Mason & Claridge, 2006). In contrast with the existing literature, our data indicates a statistically significant gender-difference in the expression of the positive dimension, with higher reported scores in females than in males (Mason & Claridge, 2006; Miettunen & Jääskeläinen, 2010). The higher expression of anhedonia described in the literature in male compared to female healthy individuals and in individuals with familial risk for psychosis (Freedman, Rock, Roberts, Cornblatt, & Erlenmeyer-Kimling, 1998) could only be observed as a trend in our sample, as the mean differences were not statistically significant.

Confirming our hypothesis, we found a statistically significant dose-dependent association of ACEs with schizotypal traits with a large effect size. Previous research had identified multiple ACEs as a major risk factor for physical and mental health conditions (Hughes et al., 2017; Riedl et al., 2020). ACEs encompass direct harm to children, such as abuse and neglect, as well as indirect impacts from their surroundings, like parental conflicts, substance abuse, or mental illness. Indeed, studies in physiology and biomolecular research have increasingly revealed how chronic childhood stress alters the development of the nervous, endocrine, and immune systems (Danese & McEwen, 2012; Pechtel & Pizzagalli, 2011). These alterations can lead to impaired cognitive, social, and emotional functioning, along with an increased allostatic load, representing chronic physiological damage. Thus, individuals with ACEs may be more susceptible to disease through differences in physiological development and the adoption and persistence of health-damaging behaviours. Maltreament is presumed to produce enduring biological change through an array of epigenetic modifications, or by gene x experience x development interactions, which are also specific to the subtype of maltreatment (Teicher, Gordon, & Nemeroff, 2022).

Prior studies have provided theoretical assumptions to explain the observed correlations between different forms of ACEs with total schizotypy scores in healthy individuals (Andorko et al., 2018; Berenbaum, Thompson, Milanek, Boden, & Bredemeier, 2008; Gawęda, Göritz, & Moritz, 2019; Goodall, Rush, Grünwald, Darling, & Tiliopoulos, 2015; Mongan, Shannon, Hanna, Boyd, & Mulholland, 2019). In our sample, emotional and sexual abuse were identified as significant predictors for the O-LIFE total scores, while experiences of neglect were not significantly associated with the O-LIFE total score. In addition, although hereditary aspects have been declared the most important factors in the transition from schizotypal disposition into the state of disease schizophrenia (Ripke et al., 2013), in our multiple regression analyses, family history of psychosis and age accounted for negligible 2.8% of the variance of schizotypy total scores. In contrast, ACEs accounted for 19.8% of this variance. This highlights the relative importance of psychosocial and environmental factors in the development and severity of schizotypal traits. Moreover, a recent study suggests that the effect of trauma in the development of interpersonal and disorganised features of schizotypy may be more apparent in the context of low genetic vulnerability for schizophrenia (Tonini et al., 2022).

At this point, it’s crucial to emphasize the comprehensive nature of the schizotypy concept, which encompasses favourable attributes. Grant coined the terms “happy schizotypes” or “benign schizotypes” to describe individuals exhibiting high levels of positive schizotypy, recurring experiences resembling psychosis, and minimal levels of disorganization and negative symptoms. Such individuals seem to obtain benefits from these traits, being characterized as creative, open-minded, imaginative, and adaptable (P. Grant & Hennig, 2020). Interestingly, the psychosis-like experiences observed in this cohort of psychologically healthy individuals are not responsible for distress, as they might be in individuals with pronounced levels of disorganization and anhedonia (C. Mohr & G. Claridge, 2015).

When discussing the relationship between ACEs and schizotypal traits, it is important to note that certain elements of the O-LIFE questionnaire encompass dissociative symptoms. This inclusion complicates the distinct delineation of these two phenomenologically different traits when relying solely on self-report questionnaires. Dissociative states are not exclusive to individuals with mental health disorders; they can also be experienced by healthy individuals and are strongly linked to childhood abuse (Draijer & Langeland, 1999; van Ijzendoorn & Schuengel, 1996). Startup observed significant correlations between the Unusual Experiences and Disorganization subscales of the O-LIFE and the Dissociative Experiences Scale, even after removing items with overlapping content (M. Startup, 1999). This persistence of correlations raises questions about the challenges in distinguishing between dissociation and schizotypy through questionnaire items, suggesting shared and underlying characteristics.

### Associations of ACEs with distinct schizotypal traits – gender-differences matter

In line with our hypothesis and the existing literature, the linear regression models investigating the relationship between various ACEs and the four schizotypal traits consistently showed an association between emotional abuse and all four schizotypal traits (Toutountzidis et al., 2022). Furthermore, our findings suggest a notable association between emotional and physical neglect and Introvertive Anhedonia. The latter finding is in line with those of Cristóbal-Narváez, who described an association between neglect and negative symptoms in young healthy adults (Cristóbal-Narváez et al., 2016), not accounting for gender differences, however. Our results demonstrated more specifically that emotional neglect showed an association with Introvertive Anhedonia in both genders, with a more pronounced effect size observed in male subjects, physical neglect exhibited an association with Introvertive Anhedonia solely in males.

Anhedonia, particularly physical anhedonia, has been identified as a significant risk factor for the transition into psychosis among high-risk patients (Bourgin et al., 2020; Schultze-Lutter, Nenadic, & Grant, 2019). From a clinical perspective, childhood neglect has been linked to symptoms of anhedonia in adolescents and young adults with depressive disorders (Cohen, McNeil, Shorey, & Temple, 2019; Wang et al., 2022). In this context, our findings on the association between childhood neglect and Introvertive Anhedonia in young, healthy adults may hold clinical significance, particularly for early monitoring of high-risk patients. Screening for ACEs, including neglect, in high-risk patients could help identify a cluster of risk factors relevant to psychosis onset. In particular, individuals with a history of neglect may be at increased risk of developing anhedonia, a significant risk factor for the transition to psychosis.

Gender differences could also be demonstrated for the association of sexual abuse with schizotypy traits. In male participants, sexual abuse was associated with Unusual Experiences, while in female subjects it was associated with Disorganisation. In a path modelling approach of data from a help seeking sample, Dizinger et al (2022) could demonstrate associations of ACEs with both positive and negative traits. These authors reported correlations between physical abuse and magical ideation, as well as emotional neglect with physical and social anhedonia in women. In men, emotional abuse predicted magical ideation, while sexual abuse was associated with perceptual aberrations (Dizinger et al., 2022). Comparing our results with those of this help seeking sample of a psychiatric early detection outpatient clinic, we found similarities and differences: in male participants, sexual abuse was associated with positive symptoms in both healthy and help seeking individuals. Interestingly, Dizinger et al found an association of neglect with anhedonia in female participants but not in male individuals, whereas in our sample, associations of neglect and anhedonia were detected in both sexes, with a stronger effect size for men than women (Dizinger et al., 2022). In line with Berenbaum et al, in our healthy community sample associations of ACEs and schizotypy traits were stronger in men than women (Berenbaum et al., 2003), contrary, in the clinical sample of Dizinger et al, which mainly included patients at high risk of psychosis suffering from schizophrenia and affective disorders, effect sizes were higher for women than men. It has been proposed that severe childhood physical and sexual abuse were associated with psychosis in female but not male patients, potentially explained by an internalizing mechanism in women coping with difficulties compared to the externalizing behaviour seen in men associated with aggression (Fisher et al., 2009). In our sample, emotional abuse exhibited a strong association with Impulsive Nonconformity in male participants and a moderate association with emotional neglect. However, no association between ACEs and Impulsive Nonconformity was found in women.

To the best of our knowledge, this study represents the first examination of detailed, gender-specific associations between ACEs and distinct schizotypal traits in healthy individuals. We hope that our findings will contribute to a deeper understanding of the complex interaction and provide insights that can inform preventive and therapeutic strategies. The gender differences are a reminder of the complexity of the relationship between two inherently complex constructs and emphasise the need for tailored interventions.

## Data Availability

All data produced in the present study are available upon reasonable request to the authors.

## Acknowledgments

This research was supported by grants from the Brain & Behavior Research Foundation and the OPO Foundation. We gratefully acknowledge their financial assistance, which made this work possible.

